# UV-C Tower for point-of-care decontamination of filtering facepiece respirators

**DOI:** 10.1101/2020.06.17.20133777

**Authors:** Badar J. Kayani, Davis T. Weaver, Vishvaan Gopalakrishnan, Eshan S. King, Emily Dolson, Nikhil Krishnan, Julia Pelesko, Michael J. Scott, Masahiro Hitomi, Jennifer L. Cadnum, Daniel F. Li, Curtis J. Donskey, Jacob G. Scott, Ian Charnas

**Author notes:** Contributed equally (Badar J. Kayani, Davis T. Weaver). **License** This work is licensed under a Creative Commons Attribution Non-Commercial Share-Alike Attribution 4.0 International License. To view a copy of this license, visit https://creativecommons.org/licenses/by-nc-sa/4.0/legalcode.

## Abstract

Filtering facepiece respirators (FFR) are critical for protecting essential personnel and limiting the spread of disease. Due to the current COVID-19 pandemic, FFR supplies are dwindling in many health systems, necessitating re-use of potentially contaminated FFR. Multiple decontamination solutions have been developed to meet this pressing need, including systems designed for bulk decontamination of FFR using vaprous hydrogen peroxide or UV-C radiation. However, the large scale on which these devices operate may not be logistically practical for small or rural health care settings or for *ad hoc* use at points-of-care. Here, we present the Synchronous UV Decontamination System (SUDS), a novel device for rapidly deployable, point-of-care decontamination using UV-C germicidal irradiation. We designed a compact, easy-to-use device capable of delivering over 2 J· cm^−2^ of UV-C radiation in one minute. We experimentally tested SUDS’ microbicidal capacity and found that it eliminates near all virus from the surface of tested FFRs, with less efficacy against pathogens embedded in the inner layers of the masks. This short decontamination time should enable care-providers to incorporate decontamination of FFR into a normal donning and doffing routine following patient encounters.

## Background and Current Challenges

Filtering facepiece respirators (FFR) are essential for protecting medical personnel and patients during outbreaks of infectious disease. In particular, the use of face shields, surgical masks, and N95 respirators are recommended for infections that may be transmitted by respiratory droplets or airborne particles.^1^ Due to the rapidly emergent nature of the novel coronavirus disease (COVID-19) and stringent requirements of proper FFR protocols, many hospitals are running dangerously low on these protective devices, to the point where they are sometimes re-used. As a result, both patients and their healthcare providers are at increased risk of contracting and spreading SARS-CoV-2, the virus responsible for COVID-19, among other pathogens.

One method of preserving our current supply of FFR is through cycles of decontamination and reuse with ultraviolet germicidal irradiation (UVGI). Substantial work has been done to evaluate the safety and efficacy of UVGI for decontamination of N95 filtering facepiece respirators (FFRs).^2-7^ Recently, UVGI has also been used to facilitate decontamination and re-use of plastic face shields.^8^ High energy UV-C rays can damage DNA and RNA, thus preventing the replication of microbes such as bacteria and viruses.^9^

Although there is no current consensus on the amount of UV radiation required to inactivate SARS-CoV-2, the UV dose required to inactivate 90% of single-stranded RNA viruses on gel media has been reported to be from 1.32 - 3.20 mJ · cm ^−2^.^2^ These estimates represent the likely dose needed to inactivate COVID-19 on face shields, while porous materials like N95 masks or surgical masks present a different challenge. While more *in vitro* studies are needed to identify the dose required for safe decontamination, literature, and subsequently governmental guidelines, suggest that a dose of at least 1 J· cm ^−2^ is required to decontaminate FFR masks prior to re-use.^10^ This relatively high required dose may make existing UVGI devices inefficient for decontamination in this context. For example, we previously described a protocol for the decontamination of FFR in biosafety cabinets available in academic laboratories. Achieving germicidal doses in these cabinets would require a minimum of 4.3 hours per-side,^7^ limiting the ease of use and throughput capacity of these devices for UVGI. These data are summarized in a recently released CDC report.^10^ UVGI and other decontamination methods are also summarized online at https://www.n95decon.org. Considering these data, we suggest that any UV-C decontamination solution should achieve a dose of at least 2 J· cm ^−2^ in a reasonable period of time.

Recently, FFR decontamination systems have been developed using vaporized hydrogen peroxide. These systems are designed to operate on large numbers of masks at a time. This approach allows for high throughput, making these systems a good solution for large hospitals. However, such large scale systems are less practical for smaller health care settings, particularly those in rural locations, or without established logistics for centralized collection and dissemination of FFR. Here, we propose a solution designed to fill this gap by enabling rapid decontamination of single masks at the point of care.

## Proposed Solution

We developed a small-footprint UV-C tower device for decontamination of FFR in a point-of-care setting (Fig S2). Our device, the Synchronous UV Decontamination System (SUDS), is small enough to be placed on a nursing station counter and can deliver more than 2 J· cm ^−2^ of UV-C irradiation to all surfaces of a mask in about a minute (Fig 2). Care providers could use our device to rapidly decontaminate their mask between patient encounters during standard handwashing protocols. Our proposed decontamination workflow using SUDS is as follows: 1) care provider doffs mask and places it in SUDS, 2) while SUDS runs, care providers can replace gloves and wash hands, 3) SUDS door opens automatically and care provider removes decontaminated mask. This workflow ensures that a care provider can continue using the same mask, which minimizes the need for re-fitting, and obviates the need for collection and dissemination. Our design and all data presented in this paper are freely available on github.

**Figure 1.**
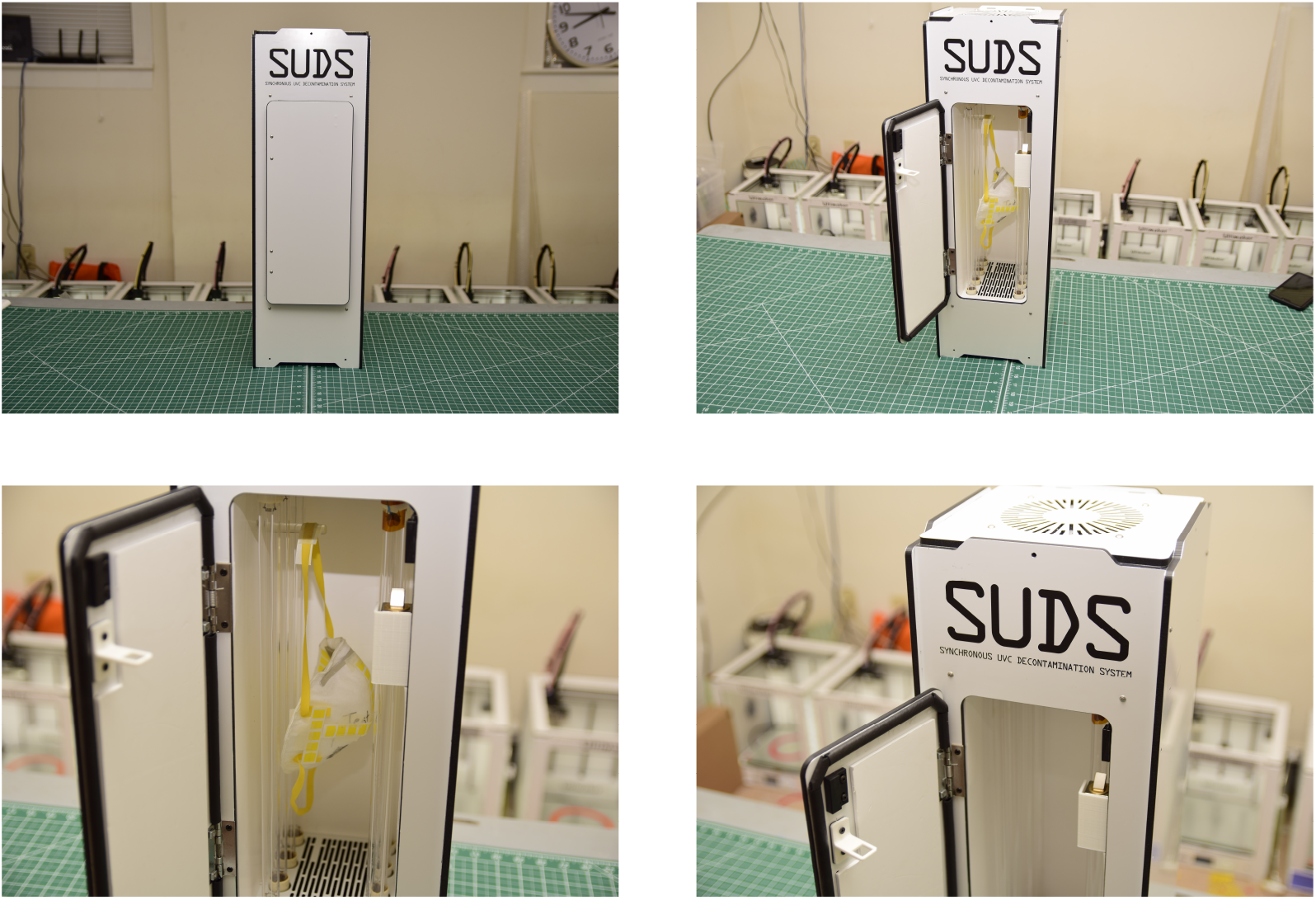
Still photos of the SUDS prototype

**Figure 2.**
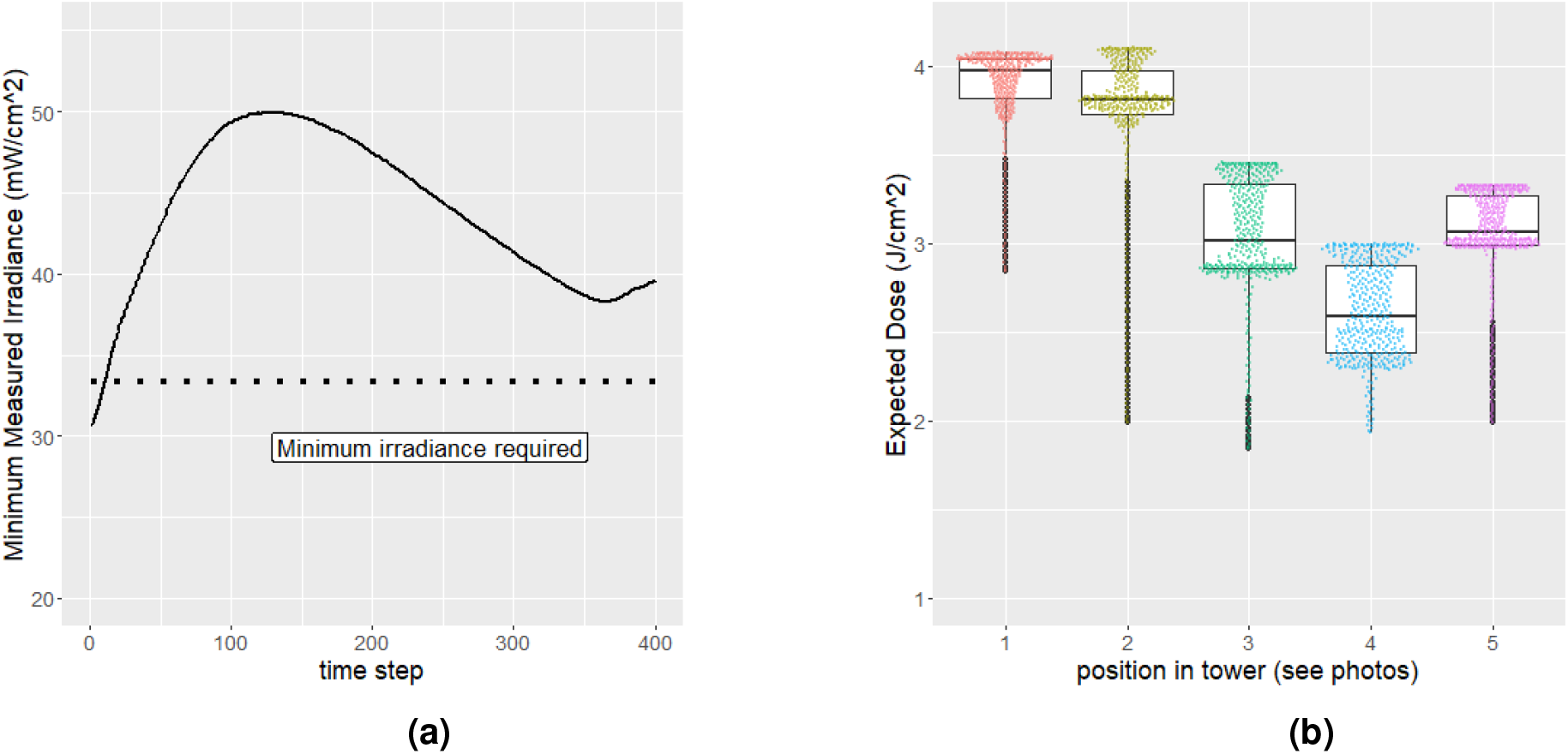
Measurements of UV-C irradiance inside the SUDS system. A. Time-series measurements of UV-C irradiance by calibrated UV fluence meter. The line corresponds to the lowest reported UV-C measurement throughout the box at a given time point. This should present the most conservative estimate of our device’s capabilities.https://www.overleaf.com/project/5e9b17ab43ff670001227d20 B: Boxplot of UV-C irradiance measurements taken at 5 different positions throughout SUDS. Measurements were converted from UV-C irradiance to expected UV-C dose after one minute of exposure.

### Design

Our design features 8 high output UV-C bulbs contained in a compact tower that combine to provide a dose of >2 J· cm ^−2^ to a single mask in about 1 minute (Fig 2). The lights are surrounded by a reflector made out of EPTFE material which reflects the UV-C light back towards the mask to allow for maximum possible dosage, and minimizes spatial heterogeneities/shadowing effects. Users hang their mask from a UV-C transparent quartz rod that suspends the mask in the center of our UV-C array. Using fused quartz (which is nearly transparent to UV-C light) ensures that there is no shadowing that could lead to areas of incomplete decontamination. We tested these novel aspects for heterogeneity and shadowing reduction by measuring the UV-C irradiance throughout the device to ensure that our array delivered at least 2 J· cm ^−2^ of UV-C, regardless of the position within the device (Fig **??**).

The device is activated upon door closure and automatically deactivates after a single cycle of decontamination is completed. The door opens automatically at the end of the cycle, allowing the user to retrieve their mask (after hand washing), limiting the risk of re-contamination.

All of the tested UV-C lamps were found to have an optimal operating temperature of around 25 - 80 °C (Fig S1). In early prototypes, we found that overheating was a serious impediment to achieving a sufficient dose of UVGI. To maintain the correct temperature and ensure adequate UV-C output, we included a thermostat which controls cooling fans in the design. The fans turn on when temperature increases beyond a set threshold. The exhaust fan intake is lined with a MERV-13 filter to prevent circulation of viral particles outside of the decontamination chamber.

The door has a magnetic sensor and a solenoid latching mechanism to keep it closed during operation. This mechanical interlock ensures that the lights will not operate when the door is open even if the solenoid latch should fail. The solenoid is actuated to release the spring loaded door after decontamination. The prototype is built out of fire-rated aluminum composite for its durability and low cost. All the main voltage electronics (including heavy ballasts and power supply) are located in the bottom of the unit to allow for better stability. The timers and relays are located in the top compartment. The entire unit runs on 110V AC supplied by a standard NEMA 5-15 plug.

### Novelty

Our device and methodology put into practice several innovative ideas that we believe represent meaningful contributions to the field of UV-C decontamination devices.

#### Speed

Most importantly, our device operates very quickly, decontaminating an N95 mask in 60 seconds. This allows a care provider to decontaminate their own FFR while they are doffing FFR and performing hand washing hygiene. Due to the speed and portability of SUDS, all hospitals, even those without the staff and experience to organize a central decontamination system, should be able to successfully decontaminate their FFR with this system. We call this “point-of-care” decontamination because it integrates seamlessly into a care provider’s workflow.

#### Highly Reflective Chamber

The inside surfaces of our device are covered with a porous EPTFE (expanded Teflon) polymer to create a highly reflective chamber to house the FFR, dramatically increasing the dose received and improving the likelihood of appropriate decontamination of FFR. Whereas aluminum (a typical commercial choice for chamber material) has a nominal UV-C reflectance of 73%, the porous EPTFE has a nominal reflectance of 97%^11^. This effect is dramatic when one considers that the light may have to reflect several times before hitting the FFR. As an illustration of this efficiency, after 10 reflections off aluminum, only 4.3% of the UV-C light will remain, whereas after 10 relfections off the porous EPTFE a much larger 73.7% of the UV-C light will remain. This reflective chamber design also ensures that both sides of the mask are decontaminated simultaneously, removing the need for users or the device to flip the mask, and ensuring both the patient-facing and health-care worker-facing sides are equally decontaminated.

#### No Shadowing

The FFR is held in place by a single hook made of fused quartz. This innovation is important to ensure the portion of the FFR’s elastomeric strap that touches the hook still receives UV-C light. Because fused quartz is 80-90% transmissive in UV-C, it ensures this portion of the strap is not significantly shadowed and does receive UV-C dose.

#### Single-Door Auto-Open Chamber

Typically, UV-C decontamination chambers contain two doors: an in-feed door where the handle and door surface are presumed to be contaminated (touched with contaminated gloves for example), and an out-feed door where the handle and door surface are presumed to be uncontaminated (touched only with decontaminated hands). While this design is effective at preventing cross-contamination, it means care must be taken to place the device in a location in the hospital where both doors will be accessible. This constraint limits where the device can be placed, potentially making it difficult to find a spot in already crowded nurse’s stations or ICU hallways. In our device, a novel use of a single auto-opening door means this device can be placed against walls or in corners, making it easier to adopt into the clinical environment.

#### Active Cooling

We found that UV bulb output was highly dependent on bulb temperature (figure S1.). By implementing active cooling in our design, we are able to deliver predictably high doses of UV-C while avoiding temperature-mediated decreases in UV irradiance.

#### UV-C Sensor Range Extension

Most commercially available UV-C sensors can measure a maximum of 20-40 mW/cm2. Because our system outputs more than these maximum irradiances, we sought to attenuate our UV-C sensors with an affordable, optically-clear plastic having partial transparency in the UV-C range. The last consideration eliminated many common plastics including polycarbonate, polystyrene, and PMMA, which block nearly all UV-C light. A material search indicated Cellophane would meet our needs, and multiple layers of cellophane were employed to ensure the received luminence was within the sensor’s operating range. We then corrected for this attenuation while processing the data. This method will be useful to any research group seeking to duplicate our results.

### Pathogen Validation

We conducted pathogen load-reduction experiments to assess the ability of SUDS to sanitize contaminated FFRs. We tested both Moldex and 3M 1860 N95 respirators under 2 conditions representing different levels of contamination. Under condition 1, samples of Clostridioides difficile (C. diff), Escherichia virus MS2 (MS2), Psueodomnas virus phi6 (Phi 6), and methicillin-resistant Staphylococcus aureus (MRSA) were suspended in an 8% mucus solution. Next, 10*μ*L of the solution was applied in triplicate to the Moldex and 3M 1860 N95 respirators, spread 10 mm, and allowed to dry. The solution was applied to the outer surface of the mask, outer edge of the mask, inner surface of the mask, and mask strap. Following inoculation, masks were treated in SUDS for 1 minute or 3 minutes. Condition 1 was designed to test the ability of SUDS to sanitize soiled or highly contaminated masks.

Under condition 2, 1 mL of the MS2 inoculum was applied to the exterior surface of each mask in triplicate and sampling was done by swabbing the exterior of the respirator. This sampling method may mimic the risk to personnel more closely than in simulation 1 as pathogens embedded within the respirator are not detected. In simulation 2, masks were treated in SUDS for 1 minute only. Control masks for both simulations were inoculated following the above protocols and left untreated. Log-reduction was calculated by comparing the SUDS treated masks to the controls. The full experimental protocol has been previously described, including more details about inoculum and viral recovery procedure^s12^.

As expected, the reduction in pathogen load varied substantially between pathogens (Fig 3). Mask location also proved to be a significant variable in pathogen load reduction. SUDS was most effective at reducing the levels of MRSA and showed moderate results in reducing the levels of Phi 6 and MS2, but fell short of significantly reducing the levels of C. diff. In addition, SUDS performed better on the inside, outside, and edge mask locations, all portions of the filtering device itself, but performed relatively worse in decontaminating the mask straps. In general, SUDS performed similarly in decontaminating the 3M and Moldex models, but performed slightly worse at decontaminating the inside surface of the 3M model relative to the Moldex model.

**Figure 3.**
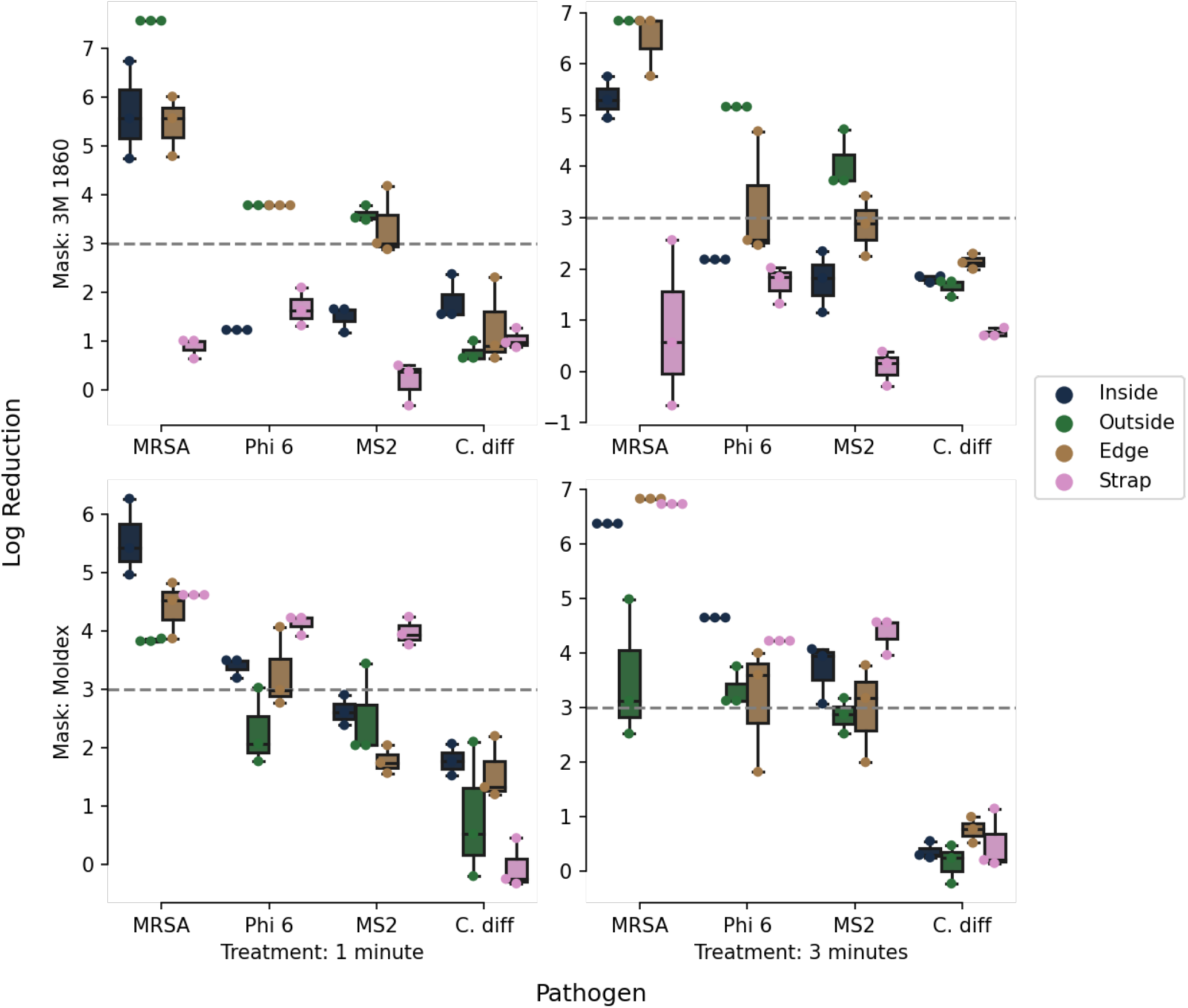
The SUDS system was highly effective at decontaminating masks soiled with MRSA, and showed varying levels of effectiveness for other pathogens. The mask straps were particularly difficult to decontaminate with UV-C for all pathogens.

Under test condition 2, which is likely more representative of the clinical use-case, almost all experiments met or exceeded a 3-log reduction in viral recovery (Fig 4). Indeed, the data that correspond to a less than 3-log reduction also exceeded the lower limit of detection, meaning that 0 infectious units were detected. The larger 500 *μ*L sample volume, which improves the lower limit of detection, met or exceeded 3-log reduction in every experiment. Thus, SUDS consistently demonstrates a 3-log reduction in MS2 under the alternative recovery protocol in test condition 2. These virology results suggest that SUDS is capable of decontaminating FFRs that have been exposed to a relatively low viral dose. It is important to note that the results from test condition 1 (Fig 3) suggest that SUDS is likely not the best instrument for the decontamination of “soiled” masks that have been exposed to infected respiratory fluids or other high-dose inoculum.

**Figure 4.**
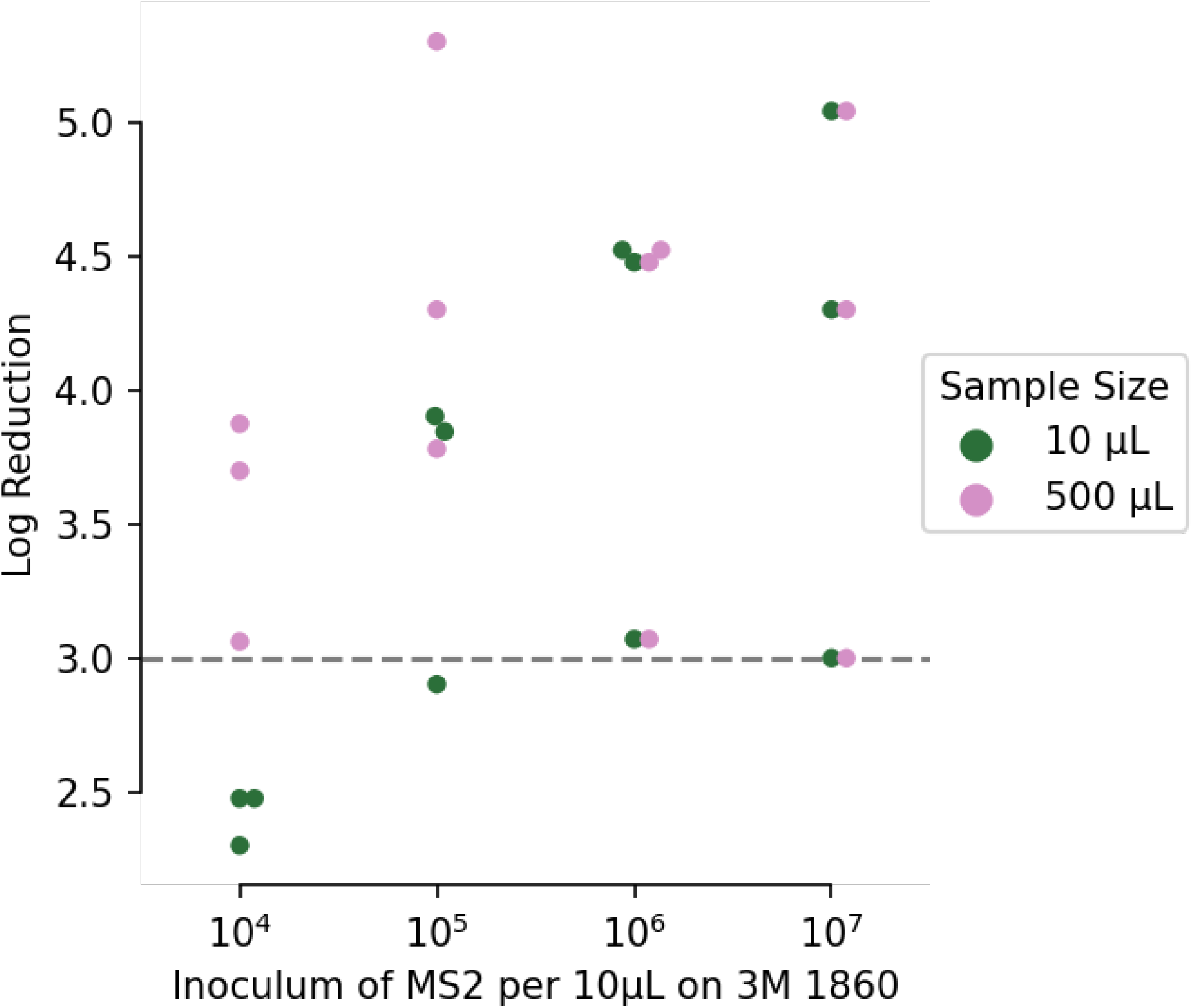
SUDS eliminates nearly all virus from the surface of the tested FFRs. When recovering virus only from the surface of the mask at the inoculums shown, SUDS achieves greater than 4 log reduction in average recovered MS2 virus.

## Discussion

Ideally, a new mask or respirator would be used for each individual to minimize the transmission of infectious diseases that are airborne or transmitted via respiratory droplets. However, crises such as the current COVID-19 pandemic can create shortages that necessitate measures to conserve FFR. Among potential methods for decontamination, previous work has suggested UVGI results in less physical deformation than bleach, microwave irradiation, and vaporized hydrogen peroxide. ^5^ Various groups have therefore begun decontaminating respiratory protective equipment themselves using UVGI and “homebrew” setups. Many of the existing solutions, however, require collection and dissemination of masks to be decontaminated in “batches”, adding logistical requirements to already busy workflows. Motivated by this, we designed the SUDS to fit directly into the workflow at the point-of-care to provide quick and easy decontamination of FFR.

According to the Food and Drug Administration “Recommendation for Sponsors Requesting EUAs for Decontamination and Bioburden Reduction Systems for Surgical Masks and Respirators During the Coronavirus Disease 2019 (COVID-19) Public Health Emergency”, SUDS achieves a Tier 3 status: Bioburden Reduction of N95 Respirators for Single Users Only to Supplement Existing CDC Reuse Recommendations.^13^ The recommendations for tier 3 status include:

1. ≥ 3-log reduction of a non-enveloped virus OR
2. ≥ 3-log reduction of two gram positive and two gram-negative vegetative bacteria OR
3. Other evidence demonstrating that the bioburden reduction system will reliably achieve >3-log reduction in non-enveloped virus or vegetative bacteria, which could include, where appropriate, published scientific literature, and scientific and engineering studies.

SUDS achieves evidence #1 and #3. As shown in figure 4, SUDS consistently demonstrates ≥ 3-log of MS2, which is a non-enveloped virus. In addition, there is some experimental evidence for the efficacy of UV-C for decontamination of FFRs, summarised by the CDC guidelines and Card et al.^7,10^

### Limitations

Despite the measures taken here to ensure adequate decontamination of FFR, following this protocol by no means guarantees complete sterilization. This device should be considered *only if* FFR *must be reused*. Virologic testing suggests that UV-C light does not adequately decontaminate the inner layers of an FFR, and is most appropriate for the sanitation of lightly contaminated, rather than soiled, FFRs. In addition, our testing suggests that the UV-C is not well suited to decontaminate the mask straps of an FFR. **Health systems electing to use SUDS or *any other UV-C decontamination protocol* should limit use to masks that have *not been exposed* to potentially infected bodily fluids**.

As discussed in the background, UV-C-mediated degradation of polymers within the respirator is another possible concern. Fit and filtration testing of the N95 respirators used in a prior experiment did not reveal any decline in filtration efficiency following UV-C exposur^e7^. Other testing also showed that, at 2 J· cm^−2^, N95 masks sustained at least 3 decontamination cycle^s14^.

## Data Availability

All code, data, and technical documents are available at https://github.com/TheoryDivision/SUDS.

## Design and Data Availability

The design and associated data is open source and publicly available.^15^

## Acknowledgements

We would like to extend our special thanks to Miguel Zubizarreta, whose generous and timely gift made this and many other COVID rapid response projects possible. We would also like to thank Ainsley Buckner, Jason Bradshaw, Umit Erol, John Kasunich, Tyler Laseter, Jim McGuffin-Cawley, Nathan McMullen, Larry Sears, Gary Wnek, and Jim Wyant for their thoughtful contributions to this work.

## Author contributions statement

This was a massive team effort with everyone contributing their specific expertise (Fig 5):

**Figure 5.**
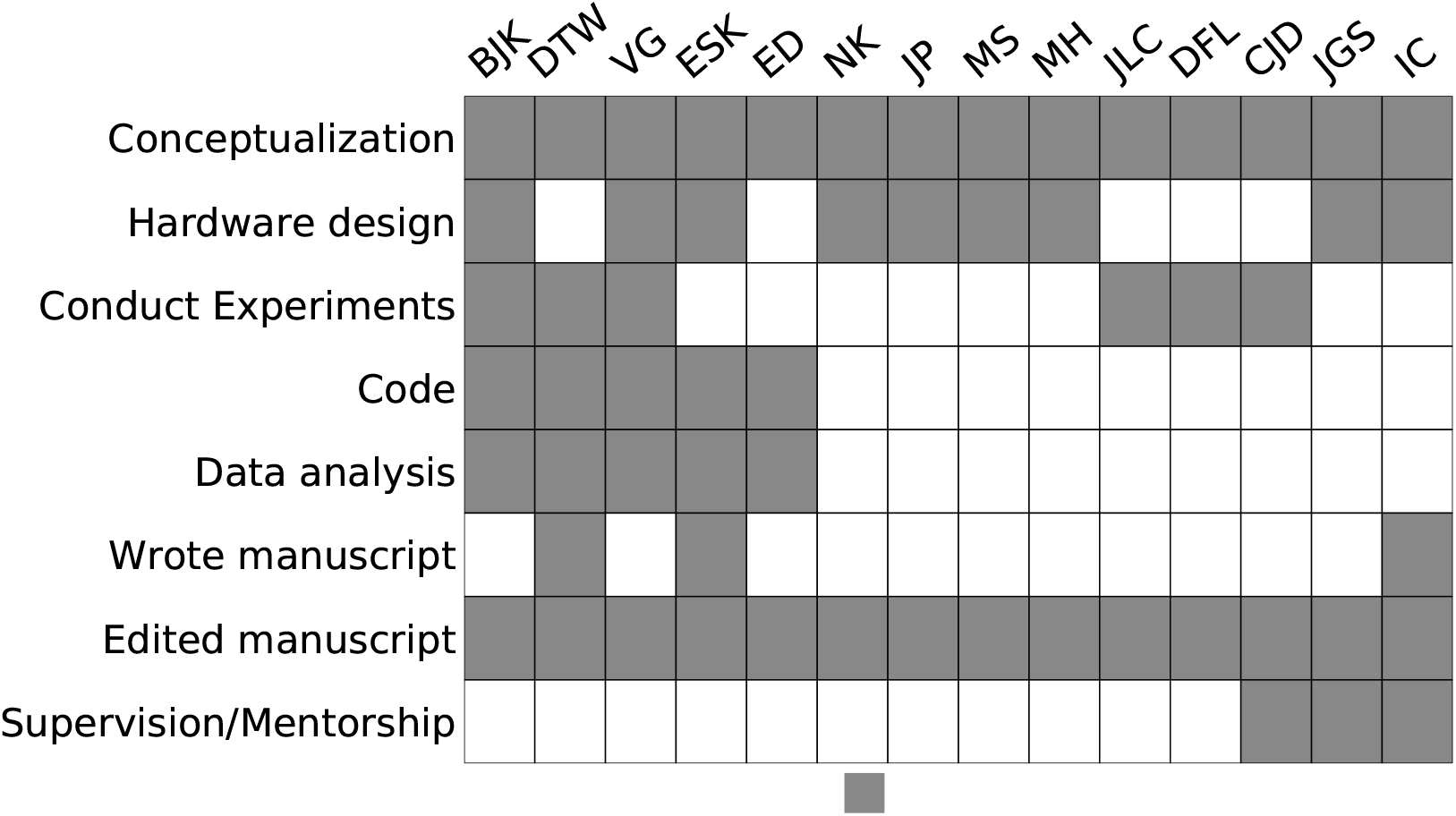
Author contributions.

## Disclosures

Jacob G. Scott and Ian Charnas are listed as the inventors on a provisional patent application for this device.

## Disclaimer

This article does not represent the official recommendation of the Cleveland Clinic or Case Western Reserve University.

## Supplemental Materials

**Figure S1.**
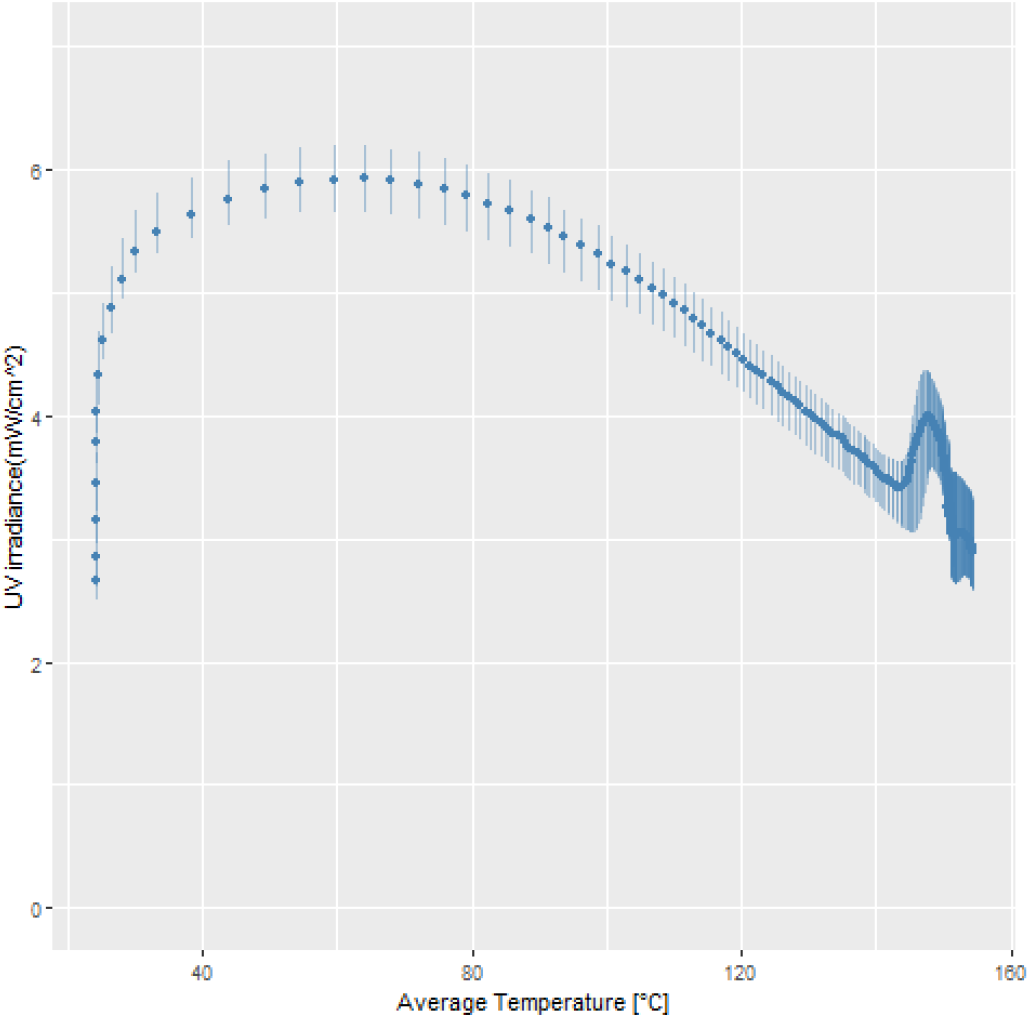
Scatterplot of bulb temperature vs. measured UV irradiance. UV-C output is heavily temperature dependent and peaks between 25 and 80 °Celsius.

**Figure S2.**
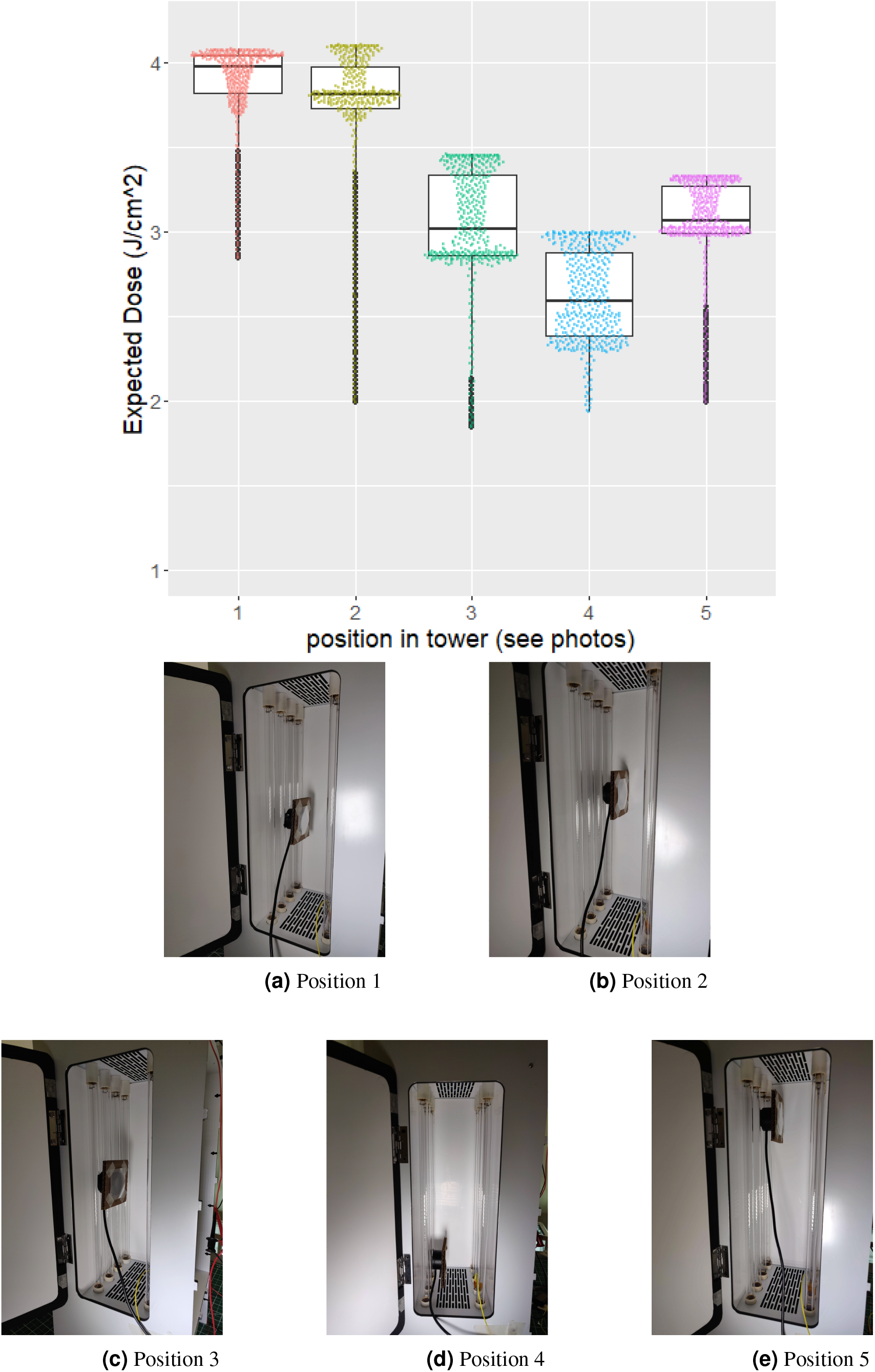
Estimated dose delivered at different positions inside the prototype. Note the UV meter was positioned at the farthest point from one side of the bulb array to generate the most conservative possible estimates of UV-C dose received by an FFR.

## Notes

### Competing Interest Statement

Ian Charnas and Jacob Scott are listed as inventors on a provisional patent application for this device

### Author Declarations

No patient data was used in this study.

